# Estimating the Risk of Outbreaks of COVID-19 Associated with Shore Leave by Merchant Ship Crews: Simulation Studies for a Case Country

**DOI:** 10.1101/2020.09.08.20190769

**Authors:** Nick Wilson, Tony Blakely, Michael Baker, Martin Eichner

## Abstract

**Aim:** We aimed to estimate the risk of COVID-19 outbreaks in a case study COVID-free destination country, associated with shore leave for merchant ship crews.

**Methods:** A stochastic version of the SEIR model CovidSIM v1.1, designed specifically for COVID-19 was utilised. It was populated with parameters for SARS-CoV-2 transmission, shipping characteristics, and plausible control measures.

**Results:** When no control interventions were in place, an outbreak of COVID-19 in our case study destination country (New Zealand; NZ) was estimated to occur after a median time of 23 days (assuming a global average for source country incidence of 2.66 new infections per 1000 population per week, a crew of 20, a voyage length of 10 days, 1 day of shore leave both in NZ and abroad, and 108 port visits by international merchant ships per week). For this example the uncertainty around when outbreaks occur is wide (an outbreak occurs with 95% probability between 1 and 124 days). The combined use of a PCR test on arrival, self-reporting of symptoms with contact tracing, and mask use during shore leave, increased this median time to 1.0 year (14 days to 5.4 years). Scenario analyses found that onboard infection chains could persist for well over 4 weeks even with crews of only 5 members.

**Conclusion:** Introduction of SARS-CoV-2 through shore leave from international shipping crews is likely, even after long voyages. The risk can be substantially mitigated by control measures such as PCR testing and mask use.

## INTRODUCTION

Historically shipping has been involved in pandemic spread globally and maritime quarantine has been used as a successful control measure e.g. in the 1918 influenza pandemic.
^1^
 Maritime quarantine has even been used successfully for preventing arrival of the 2009 influenza pandemic in some island jurisdictions.
^2^

The COVID-19 pandemic has also had an impact on maritime vessels during 2020, along with spread to people on shore. On the *Diamond Princess* 19% of the passengers and crew became positive with the pandemic virus (SARS-CoV-2) and there was spread to Japanese responders on shore.
^3^
 Similarly, on the *Grand Princess*, 17% of those tested had positive results.
^3^
 On a much smaller cruise ship with 217 passengers and crew onboard, 59% were reported to be test-positive.
^4^
 On a fishing vessel, 85% (104/122) of the crew were infected.
^5^
 In terms of merchant vessels, an outbreak on a container ship was reported as infecting 23% (5/22) of the crew.
^6^
 Other such outbreaks have been detailed in media reporting (referred to in a review
^7^
).

In response to the COVID-19 pandemic, border controls have been widely used to limit pandemic spread. Such border controls are particularly relevant for two types of pandemic control strategy: (i) the exclusion strategy as successfully practiced by ten Pacific island nations e.g. Samoa and Tonga;
^8^
 and (ii) the elimination strategy as used by New Zealand,
^9^
 and possibly other jurisdictions, e.g. Mainland China, Taiwan, Fiji and five states/territories in Australia.

Some of these jurisdictions have completely prohibited maritime vessels arriving at their sea ports from countries which are not COVID-19-free (e.g. the Marshall Islands have prohibited such incoming ships
^8^
). But time periods are also used e.g. a minimum of 14 days at sea before being allowed to enter the Marshall Islands,
^8^
 or 14 days plus a negative PRC test for New Zealand.
^10^
 There is also the standard international requirement for pratique whereby any “illness during the voyage” must be notified to health authorities at the destination port.
^11^

Given this background we aimed to expand on previous modelling work (for air transport spread of COVID-19^12^) to determine the risk of merchant ships being the source of COVID-19 outbreaks in an otherwise COVID-19 free country.

## METHODS

### Model design and parameters for SARS-CoV-2 and COVID-19

We used a stochastic SEIR type model with key compartments for: susceptible [S], exposed [E], infected [I], and recovered/removed [R]. The model is a stochastic version of CovidSIM which was developed specifically for COVID-19 (http://covidsim.eu; version 1.1). Work has been produced from previous versions of this model,^
12 13 14^ and in two places we detail the relevant equations and their stochastic treatment.^15 16^ The model was built in Pascal and the computer code is available on request from the senior author (ME).

100 million simulations were run for each set of parameter values. Such a large number of simulations was necessary due to the very high probability of zero infected crew members boarding a departing merchant ship given the low assumed incidence of infection (see below). The overall framework for the processes modelled is shown in Figure 1. The parameters were based on available publications and best estimates used in the published modelling work on COVID-19 (as known to us on 27 August 2020). We assumed that 71% of infected COVID-19 cases develop clearly detectable symptoms (Table 1). Another assumption was the contagiousness in terms of the effective reproduction number (R_eff_) which was 3.0 among crew members on board of the ship and 2.5 in the destination country (Table 1).

**Figure 1:**
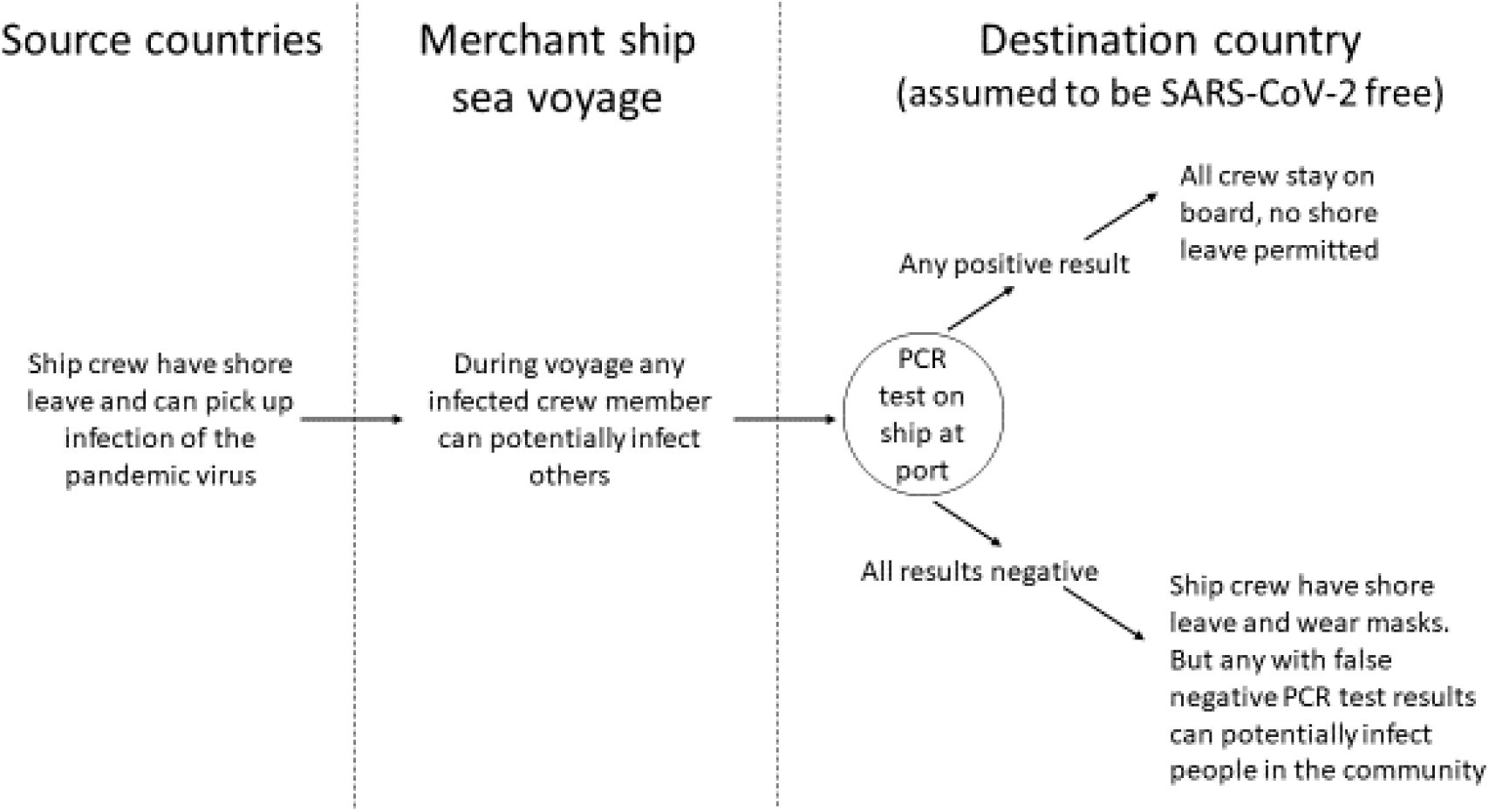
Flow diagram of the assumed movements of merchant ship crews in the model including interventions (simplified and not showing all control measures e.g. the seeking of medical attention when symptomatic in the destination country and the associated isolation of identified cases and contact tracing)

**Table 1:** Input parameters used for modelling the potential spread of SARS-CoV-2 infections via merchant shipping with the stochastic version of CovidSIM (v1.1)^17,18,19,20,21,22,23,24,25,26-28,29,30,31^

| Parameter | Value/s used | Further details for parameter inputs into the modelling |
| --- | --- | --- |
| Incidence of SARS-CoV-2 infection (using the global average) | Daily incidence = 0.00038 (i.e. 2.66 infections per 1000 population per week) | We estimated the incidence of new infections globally for 15 August 2020 using the following data and assumptions:<br>For the initial estimate of the numerator we used the global reporting to WHO of new laboratory-confirmed cases of SARS-CoV-2 infection on 15 August 2020 ($n=294,237$ new cases).<sup>17</sup><br>For the denominator we used the UN global population estimate for 2020 (7,794,799,000).<sup>18</sup><br>To adjust for under-estimating of actual infections (compared to reporting of cases) we used the estimate of a 10-fold difference between reported cases and infections based on sero-surveys by Havers *et al.* for the USA (with this 10 fold factor still probably being an under-estimate).<sup>19</sup><br>We assumed that prior to the ship leaving for the destination country, the crew members have 1 day of shore leave during which they can pick up the infection at the given probability. |
| Percent of infections that are asymptomatic | 29% (50% in scenario analysis) | We used the estimate from a very large Spanish survey of 61,075 participants.<sup>20</sup> It found the proportion of individuals with a positive test who were asymptomatic was 32.7% (30.2–35.4) for the point-of-care test and 28.5% (25.6–31.6), for the immunoassay. Given the immunoassay is likely to be more accurate than the point-of-care test, we used the 28.5% result. This result is similar to that for a working-age adult population i.e. a cohort of health care workers in the UK at 27% of all infections being asymptomatic.<sup>21</sup> |
| Latency period | 5 days | We used the best estimate from CDC in May 2020 of a mean of 6 days until symptoms (i.e. the latency period plus the prodromal period).<sup>22</sup> We used a standard deviation (SD) of 25% (1.25 days) (calculated using 16 stages; Erlang distribution). |
| Prodromal period | 1 day | There is, as yet, insufficient information on this prodromal period for COVID-19, so we used an assumed value for influenza (SD = 25%; 0.25 days, Erlang distribution). |
| Symptomatic period | 10 days (split into 2 periods of 5 days each) | The WHO-China Joint Mission report stated that “the median time from onset to clinical recovery for mild cases is approximately 2 weeks and is 3-6 weeks for patients with severe or critical disease”.<sup>23</sup> But given that mild cases may have been missed in this particular assessment, we used a slightly shorter total time period of 10 days (SD = 25%; 2.5 days, Erlang distribution). |
| **Contagiousness** |  |  |
| Relative contagiousness in the prodromal period | 100% | We used the best estimate from CDC in May 2020 of infectiousness of asymptomatic individuals relative to symptomatic individuals of 100%.<sup>22</sup> |
| Contagiousness after the prodromal period | 100% and 50% | In the first five days of symptoms, cases were considered to be fully contagious. In the second five-day period, this was assumed to be at 50%. The latter figure is still uncertain, but is broadly consistent with one study on changing viral load.<sup>24</sup> |
| Effective reproduction number ($R_{\text{eff}}$) on board the ship | 3.0 (4.0 in a scenario analysis) | The enclosed nature of the ship environment (and shared sleeping quarters in smaller vessels of under 3000 gross tonnage), would favour disease spread and so we used a higher value than for the community (see below). Noting the fishing boat outbreak (detailed in the *Introduction*) where 85% of the crew became infected,<sup>5</sup> we also used a higher value ($R_{\text{eff}} = 4.0$) in a scenario analysis. We assumed no routine mask use on the ship or specific additional physical distancing behaviours by the crew. |
| $R_{eff}$ in the destination country | 2.5<br>(2.0 in a scenario analysis) | We used the best estimate from the CDC of 2.5 for community transmission.<sup>25</sup> We assumed a COVID-19-free destination country such as New Zealand (NZ) where human social behaviour in the elimination period (May to July 2020 in NZ) was fairly similar to the pre-COVID-19 state (i.e. relatively little additional physical distancing, normal occurrence of large social events and no routine mask use by the great majority of the population). Nevertheless, we also considered a value of 2.0 in a scenario analysis. We assumed a population with no specific immunity to SARS-CoV-2. |
| Super-spreading in the destination country | Just in a scenario analysis | Given some evidence for super-spreading phenomena with this pandemic virus,<sup>26-28</sup> we also considered a scenario where in NZ just 10% of the cases generated 10 times as many secondary cases as the other cases. |
| **Shipping-related parameters** |  |  |
| Merchant ship visits to the destination country | 108 per week | In the last three quarters of 2019 and the first quarter of 2020 there were 5600 merchant ship port visits in NZ by vessels originating in overseas ports (counting each port visit separately where multiple ports were visited).<sup>29</sup> This is 108 port visits per week for such vessels. These vessels include bulk carriers, container ships, reefers, tankers, vehicle carriers and a range of other types of cargo vessels. |
| Voyage length | 10 days<br>(scenario analyses ranging from 1 to 30 days) | We calculated merchant ship travel times using a specific website for travel times between sea ports (http://ports.com/) and using a typical travel speed of 24 knots (44 km per hour). This gave the shortest trip to NZ (Sydney to Auckland) at 1330 nautical miles (nm) [2463 km] taking 2.3 days at sea. It gave the longest possible trip to NZ (Montreal to Auckland) at 17,100 nm taking 29.7 days at sea. Also, it gave the trip from the world's busiest container port (Singapore) to Auckland at 5828 nm at 10.1 days at sea. It gave the trip from the busiest European container port (Rotterdam) to Auckland at 14,569 nm at 25.4 days at sea. Given the complexities we did not consider port calls and shore leave on route between the original departure point and the first NZ port of call. Also we note that delays can also result in slower voyages than these (e.g. from storms, port congestion etc.). |
| Crew size | 20<br>(scenario analyses: 5, 10, 30) | This value varies for the type of merchant vessel, but we used a figure of 20 which is mid-range for the crew size of a container ship (range 10 to 30 crew).<sup>30</sup> A wider range of values was used in scenario analyses. |
| Duration of shore leave | 1 day<br>(scenario analyses: 2, 3) | We analysed Port of Auckland data (the port in NZ's largest city) for the 140 merchant ship visits detailed on their website for 20 August 2020. This indicated a median stay in this port of 1 day (range 0.3 days to 6 days).<sup>31</sup> However, 31% of these international merchant ships had most recently come from another NZ port prior to the Port of Auckland. |

### Shore leave in the destination country

We selected New Zealand as a case study destination country as it has previously achieved elimination of community transmission of SARS-CoV-2^9^ and appears to be successfully controlling an outbreak (probably arising from a border control failure) in one region in September 2020. Upon arrival in this destination country, we used a period of shore leave by all the crews of one day (the median time ships are in port based on Ports of Auckland data in as, the port in New Zealand’s largest city).

### Potential control measures

Are detailed in Table 2 and Figure 1 and included a PCR test on all the crews on arrival and mask use by the crews during shore leave. If any crew member tested positive then the shore leave for all that particular crew was assumed to be prohibited and therefore there was no risk of any community outbreak. If a crew member on shore leave developed and self-reported symptoms and then tested positive, this case would be isolated and this could also trigger contact tracing which was assumed to identify 80% of the infected contacts within 48 hours. Identified contacts would be isolated after a delay of one or two days.

### Ongoing infection transmission in the destination country

Untraced secondary cases who were infected by crew members in the destination country, and tertiary cases who were infected by traced secondary cases before they were isolated, were assumed to roam freely for the full length of their infectious period and to potentially trigger outbreaks in the community.

### Control measures assumptions

The full details on the considered control measures are given in Table 2.

**Table 2:** Control measures used and their estimated efficacy^32,33,34,35,36,37,38^

| Control measure | Key value | Comment |
| --- | --- | --- |
| Pratique | Not considered as PCR testing more accurate | Although some cases of SARS-CoV-2 infection will be asymptomatic (see above) and others fairly mild, it is likely that a proportion of onboard outbreaks of COVID-19 would come to the awareness of the ship's captain. A small proportion of cases would also become seriously ill requiring immediate treatment and potentially the diversion of the ship to a nearby port (or removal of a case by helicopter). The |
|  |  | captain would be then legally required to alert health authorities in the destination port as part of pratique. On the other hand if a captain knows that the crew are in particular need of shore leave, then such information about onboard outbreaks might not always be divulged. The captain may also discount any outbreaks of respiratory illness as being due to other causes and to have been resolved at the time of arrival. Hence we assumed that port authorities should place little emphasis on pratique as a control process and should require PCR testing of all crew wanting shore leave (as outlined below). |
| Compulsory PCR test on arrival of all crew | Variable sensitivity based on time since infection | As per our previous modelling work,<sup>12</sup> we used the results of a study<sup>32</sup> which fitted a Bayesian hierarchical logistic regression model for test sensitivity. This meant for example, at day 4 after infection, 67% of test results were false negatives (95%CI: 27% to 94%). This decreased to 20% (95%CI: 12% to 30%) on day 8 and then increased after this e.g. up to 66% (95%CI: 54% to 77%) on day 21. For cases who already recovered before their PCR test, we use the final value reported by Kucirka *et al.* (i.e. 34% sensitivity).<br>We assumed all crew would request shore leave and that port health authorities would prioritise the PCR testing of seafarers immediately on arrival to allow for a day of shore leave. E.g. we note that as per some airports, PCR test results can be obtained within a few hours.<sup>33</sup> We also note imminent access to faster testing e.g. FDA approval of a 15 minute test (albeit with different sensitivity and specificity from the PCR test).<sup>34</sup> |
| Mandatory mask use by the crews during shore leave | 85% efficacy but only two-thirds (66.7%) adherence (and one third adherence in scenario analysis) | We used the efficacy value of 85% from a systematic review and meta-analysis (n=2647; adjusted odds ratio = 0.15, 95%CI: 0.07 to 0.34).<sup>35</sup> Adherence to mask use in social settings in NZ (where local citizens are not typically using masks except on public transport where it was mandated in August 2020) was considered likely to be suboptimal at two thirds. In a scenario analysis we set adherence to mask use at one third (33.3%). |
| Self-reporting of crew members whose sickness starts shortly before or during shore leave (i.e. they are among the 71% of infected individuals who become symptomatic) | 50% (self-reporting, occurring on average 1 day after symptom onset) | We used the same estimated value as in our previous Australia to NZ air travel study.<sup>12</sup> Such reporting can trigger contact tracing amongst the public in the destination country and therefore lower the risk of an outbreak (see next item). But due to the complexities we do not consider backward contact tracing among the crew. Of note is that routinely in NZ, 39.5% of people with “fever and cough” symptoms seek medical attention, as reported by the NZ Flutracking surveillance system.<sup>36</sup> This is very similar to international estimates for people with influenza who seeking medical attention at 40% e.g. as used in other modelling.<sup>13</sup> |
| Contact tracing if crew members develop symptoms in NZ, seek medical attention and are confirmed by PCR (see above) | 80% of infected contacts are traced and isolated within 48 hours | We used performance data for the cluster of cases in Auckland in August 2020 where the official estimate was 80% of contacts contacted within 48 hours (as reported by the Prime Minister).<sup>37</sup> We divided this into 60% within the first 24 hours and 20% in the next 24 hours. Of note is that variable performance for contact tracing has been reported for NZ at other times in August 2020, with 86% of contacts traced in 48 hours at one point.<sup>38</sup> |

## RESULTS

The results of the stochastic simulations indicate that if no pandemic-related maritime controls were in place, the COVID-19-free destination country (New Zealand) would quickly experience an outbreak attributed to ship arrivals. That is an outbreak after a median duration of 0.064 years (23 days) which is equivalent to a total of 355 port visits and 7100 total days of shore leave (for international 20 crew members per vessel, and one day of shore leave per port; Table 3). There is high uncertainty however, with 95% of outbreaks likely to occur between 0.0023 and 0.34 year (i.e. 1 to 124 days; Table 3).

**Table 3:** Results of the simulations without interventions and with multi-layered interventions (for a base case of 10 days at sea and 108 merchant ship visits per week, 20 crew per ship, one day of shore leave each per port visit in New Zealand (NZ), 100 million stochastic simulations were run for each set of parameters).

| PCR test upon entry | Wearing face masks when on shore leave (by the crew) | Self-reporting of symptoms (when in NZ) | Contact tracing for self-reported cases | Median duration until outbreak in NZ (years) | 95% of outbreaks are likely to occur in this time interval [years] |
| --- | --- | --- | --- | --- | --- |
| no | no | no | no | 0.06 | 0.002 – 0.34 |
| yes | no | no | no | 0.46 | 0.017 – 2.47 |
| yes | yes | no | no | 1.00 | 0.037 – 5.53 |
| yes | yes | yes | no | 1.02 | 0.037 – 5.41 |
| yes | yes | yes | yes | 1.02 | 0.037 – 5.43 |

The median time to an outbreak would be markedly increased by obligatory PCR testing of crew members before shore leave is permitted i.e. up to 0.46 years (168 days) or after a total of 2592 port visits. Even further reduction of risk occurs when requiring face mask use during shore leave (increased median time to 1.00 years). But relatively little extra gain in risk reduction occurs from any sick crew on shore leave self-reporting symptoms and from the associated contact tracing (Table 3). Using the base case value of R_eff_ = 2.5 in New Zealand, a single untraced infection in the community leads to an outbreak in 88.2% of cases (78.5% for R_eff_ = 2.0). When we considered super-spreading events in the community in a scenario analysis the outbreak probability per person was actually reduced to 57.4%. This is because allowing for super-spreading events means that a smaller proportion of infected crew members transmit infection, even though those that do will typically infect more people (assuming the same overall value of R_eff_).

In scenario analyses, a smaller crew size reduced the outbreak risk (e.g. the median time to an outbreak would be 3.8 years for ships with a crew size of five; Table 4). The risk of outbreaks was also lower when making assumptions around lower contagiousness in the destination country (i.e. R_eff_ lowered to 2.0). The risk remained basically unchanged if contagiousness on the ship was assumed to be higher (i.e. R_eff_ increased to 4.0). Increasing the shore leave to either two or three days increased the risk of an outbreak (i.e. it reduced the median waiting time). If super-spreading events were considered in the destination country, this led to the same average number of untraced infections caused by crew members in New Zealand, but as each one of them had a lower risk of leading to an outbreak (see above), the overall outbreak risk was lower than in the baseline study.

**Table 4:** Results of the scenario analyses for 108 merchant ship visits per week and the full set of interventions taking place (see last line of Table 3) with 100 million stochastic simulations run for each set of parameters (for further information, see text and Table 2).

| Scenario (compared to base case) | Median duration until outbreak in NZ (years) | 95% of outbreaks are likely to occur in this time interval [years] |
| --- | --- | --- |
| Base case with all interventions (for comparison purposes) | 1.02 | 0.037 – 5.43 |
| 5 crew members instead of 20 | 3.81 | 0.139 – 20.27 |
| 10 crew members instead of 20 | 2.02 | 0.074 – 10.74 |
| 30 crew members instead of 20 | 0.68 | 0.025 – 3.63 |
| 2 days of shore leave instead of 1 | 0.28 | 0.010 – 1.51 |
| 3 days of shore leave instead of 1 | 0.14 | 0.005 – 0.74 |
| $R_{\text{eff}}$ in NZ is 2.0 instead of 2.5 | 1.38 | 0.050 – 7.34 |
| $R_{\text{eff}}$ on board the ship is 4.0 instead of 3.0 | 1.07 | 0.039 – 5.68 |
| Super-spreading events can occur in NZ | 1.61 | 0.059 – 8.54 |
| 50% of infections are asymptomatic instead of 29% | 1.01 | 0.037 – 5.36 |
| Mask use adherence during shore leave is one third (33%) instead of two thirds (67%) | 0.64 | 0.023 – 3.41 |

Figure 2 shows that voyage duration is a key determinant of outbreak risk in the destination country and this risk is especially high for short voyages of under a week (i.e. when infected crews taking shore leave may still be PCR test negative). This Figure also shows that it takes a long time for the onboard epidemic to “burn out” and that the outbreak risk in the destination country (when there are no controls) only starts to decline after a voyage time of three weeks, and even then declines quite slowly (Figure 2a). For a crew size of 20 the risk of community outbreaks is still increasing after four weeks of voyaging if no controls are used (Figure 2c). Interestingly, if PCR tests are implemented, the effect of longer travel durations generates results that are the inverse: the more the infection can spread on board, the more likely it will be detected. As none of the crew members is assumed to be allowed to go to shore if one is found positive, the probability that infected people entering the destination country decreases with the number of infected people on board. Adding additional interventions like wearing masks, self-reporting symptoms and doing contact tracing further improves the results, but the main effect is obtained by PCR entry screening. With the full set of interventions the median time to an outbreak increased up to 25 years (Figure 2b and 2c).

**Figure 2:**
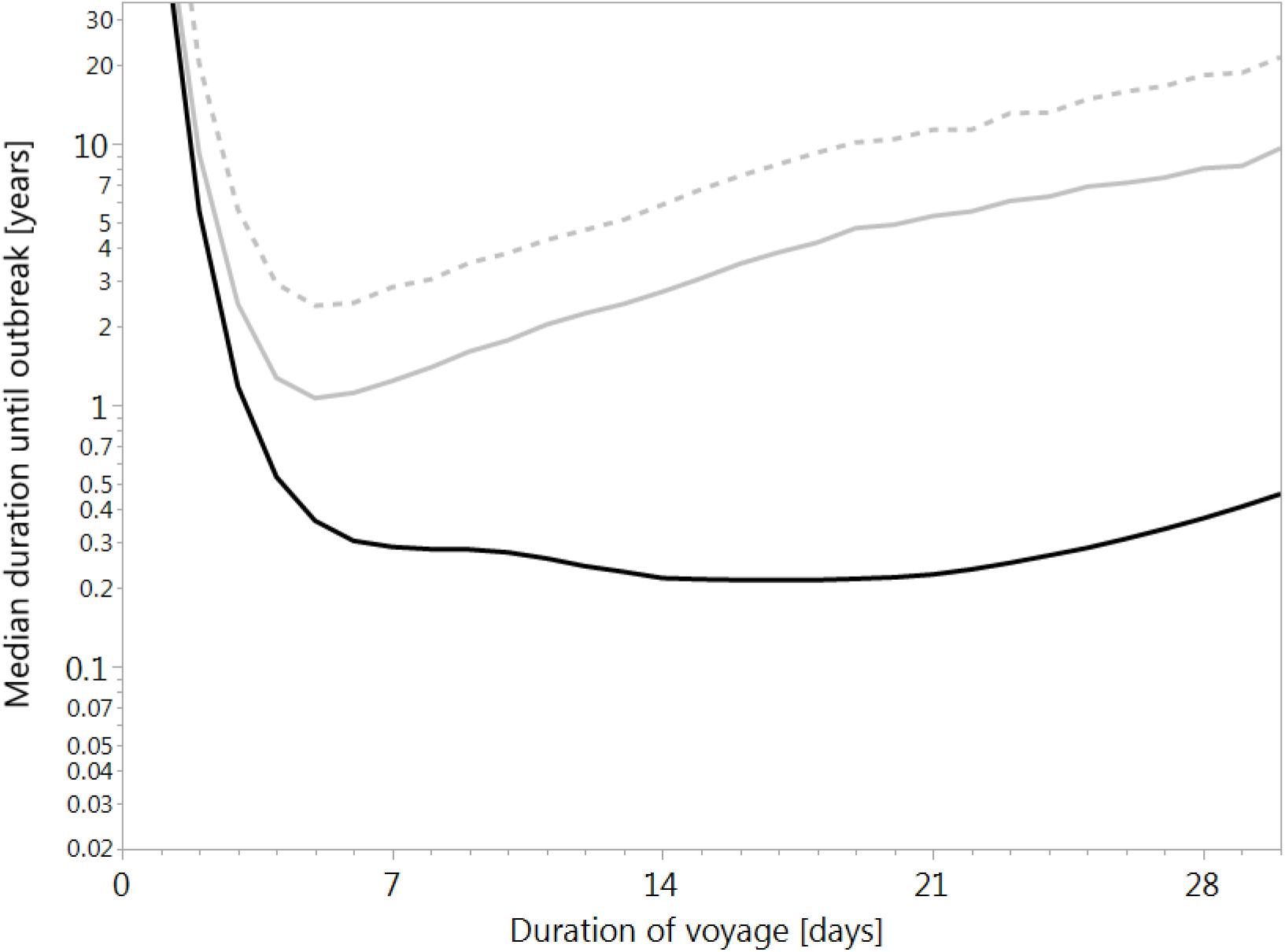
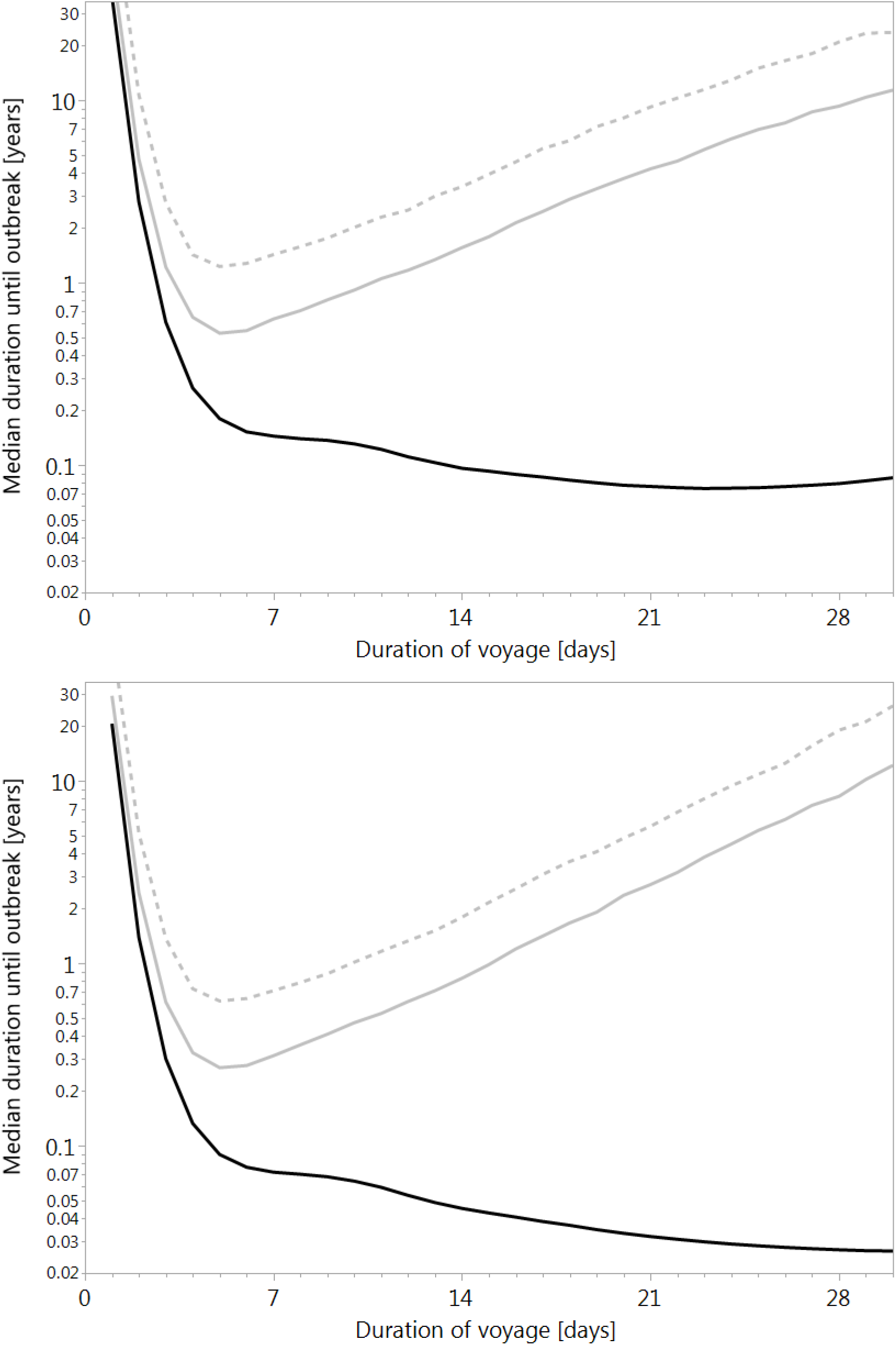
Median duration [log-scale in years] until a COVID-19 pandemic outbreak occurs in the destination country because of merchant ship crews taking shore leave. (a) 5, (b) 10, (c) 20 crew members per ship; 108 cargo ships arrive each week. In the country of origin, each member can pick up the infection at a rate of 0.00038 per day. Infections spread on board with an effective reproduction number R_eff_ of 3.0 and in NZ with R_eff_ of 2.5. Note that a voyage duration of 1 day is not applicable to NZ. Full black curves: no interventions are taken; full grey curves: all crew members are prevented from entering the country if one of them is PCR positive upon arrival; dotted grey curves: full set of interventions as outlined in Table 3. For each combination of crew size and voyage duration, 100 million voyages were simulated.

## DISCUSSION

### Main findings

The results of this modelling study suggest it would only be a matter of a few weeks (specifically for the base case around 23 days for a total 355 port visits and 7100 days of shore leave) before crew from international trading maritime vessels would trigger COVID-19 pandemic outbreaks in the destination country. Fortunately, however, the risk of such outbreaks can be substantially reduced with available interventions, especially PCR testing before leaving the vessel and use of masks by the crew during shore leave. Of particular note is that even small five person crews will contribute to a risk after voyages of several weeks and this risk does not start to decline until three weeks (and even then the decline is slow).

It is likely the results for our case study country (New Zealand) are generalisable to most countries that have sea ports and maritime trade. Nevertheless, the risk could be somewhat less for some nations on a per population or per GDP basis because New Zealand’s economy is particularly trade orientated and especially sea trade orientated. That is, it has no international trade by land routes and only a small proportion by air cargo. With a population of 5 million New Zealand has 1120 port visits from vessels with an international origin per million population per year.

### Study strengths and limitations

This appears to be the first modelling study to explore the risk of COVID-19 outbreaks arising from shore leave of maritime ship crews (based on our search of PubMed and preprint sites in August 2020). Another strength is that the work builds on an established model that has been used to also study air transport and other aspects of SARS-CoV-2 transmission (see 
*Methods*
).

But as with all modelling there are important limitations. Some of these relate to parameters, with a particularly critical one being the daily incidence of SARS-CoV-2 infection in the source country that the ship leaves from. We just used a global average for this incidence to account for the diverse maritime trading patterns that New Zealand has and also because the crews are also internationally diverse (often flying in from another country just prior to the ship’s departure). Nevertheless, there are likely to be highly variable risks by source country and countries that the crew come from.

Another example of parameter limitations are the R_eff_ onboard such vessels and also the R_eff_ for shore leave by crew. The former is likely to vary by different designs of merchant vessels (container ships vs. tankers vs. bulk carriers etc.) and also by size (e.g. it is likely that in vessels of under 3000 gross tonnage the crew are in shared sleeping rooms). However, we did not have sufficient data to model such heterogeneity. We also didn’t account for potential immunity amongst crew from past exposure to the SARS-CoV-2 pandemic virus internationally, which is bound to increase over time. Given the complexities we did not consider port calls and shore leave on route between the original departure point and the first New Zealand port of call. However, such port calls (if shore leave is taken by at least some of the crew members) could be reconceptualised as the new starting point for the voyage. We also did not model risk of transmission to port workers who might go onto arriving ships (eg, pilots and health workers conducting PCR tests), on the assumption that they would take appropriate precautions with physical distancing and use of personal protective equipment.

### Possible implications for future research and policy

Future research is needed to replicate this study, e.g. using simulation models with a different structure and for a wider range of destination countries. Research could also explore the acceptability and adherence to mask use by crews on shore leave in different settings.

As detailed above, the results in Tables 2 and 3 might make some health authorities decide that the risk of allowing shore leave for crew is tolerable with control interventions such as PCR and masks in place. But for small low-income island states (e.g. the 10 nations in the Pacific that were COVID-19-free in September 2020) the risk might still be considered too high, especially if they have limited surveillance and outbreak control capacity. In these states, either all shore leave could be denied (i.e. cargo movement is performed without the crew leaving the vessel), or the ships which recently visited countries where COVID-19 transmission is occurring are completely prohibited (e.g. until a vaccine against COVID-19 is available).

### Conclusions

Using simulations, we estimated the risk of COVID-19 outbreaks in COVID-19-free settings as a result of merchant ship crews taking shore leave. Our results can inform policy-maker decisions about regulations regarding shore leave for crews and the use of various control measures such as PCR testing and mask use to minimise the risks if shore leave is permitted.

## Data Availability

The computer code used in the model (Pascal) is available on request from the senior author.

